# Physical performance and risk of cardiovascular and all-cause mortality in the United States: The Reasons for Geographic and Racial Differences in Stroke (REGARDS) study

**DOI:** 10.64898/2026.03.30.26349789

**Authors:** Abu Abdullah Mohammad Hanif, Parag Goyal, Lisandro D. Colantonio, Monika M. Safford, Ene M. Enogela, Ro-Jay Reid, Mojisola Fasokun, Philip Akinyelure, C. Barrett Bowling, Hugo G Quezada-Pinedo, Madeline R. Sterling, Emily B. Levitan

**Affiliations:** Department of Epidemiology, University of Alabama at Birmingham, Birmingham, AL; Division of General Internal Medicine, Weill Cornell Medicine, New York, NY; Department of Medicine, Duke University School of Medicine, Durham NC; Durham Veterans Affairs Geriatric Research Education and Clinical Center, Durham Veterans Affairs Medical Center (VAMC), Durham, NC; Department of Population Health Sciences, Duke University School of Medicine, Durham NC, USA

**Author notes:** Corresponding author: Emily B. Levitan, ScD, Department of Epidemiology, RPHB 220, University of Alabama at Birmingham, Birmingham, AL 35294-0022, USA.

**Keywords:** Physical Performance, mortality, CVD, all-cause, REGARDS, US

## Abstract

**Background:** Poor physical performance, measured by gait speed and chair stands, is associated with mortality; associations may differ by history of cardiovascular disease (CVD).

**Methods:** Among 14,137 REasons for Geographic And Racial Differences in Stroke (REGARDS) study participants, gait speed and chair stand times (2013–2016) were categorized into quartiles and a fifth category with those who were unable to complete the test. Associations with adjudicated CVD and all-cause mortality through 2020 were examined among participants with and without history of CVD.

**Results:** Average age was 72.5 ± 8.5 years. Among participants without history of CVD, those in slowest vs. highest gait speed quartile had HRs of 2.01 (95% CI 1.18–3.43) for CVD and 1.66 (1.33–2.07) for all-cause mortality; among those unable to complete the test, HRs were 2.37 (1.12–5.03) for CVD and 2.33 (1.72–3.17) for all-cause mortality. Among participants with history of CVD, slowest gait speed quartile had HRs of 1.28 (0.96–1.72) for CVD and 1.72 (1.45–2.04) for all-cause mortality; HR among those unable to complete the test were 1.87 (1.29–2.70) for CVD and 2.74 (2.22–3.38) for all-cause mortality (p-interaction between with and without history of CVD <0.05). Inability to complete chair stand test was associated with higher mortality in both groups.

**Conclusions:** Poor physical performance was associated with greater CVD-related and all-cause mortality among both individuals with and without a history of CVD, with the highest risks observed among those who were unable to the assessments.

## Introduction

Physical function is an important indicator of health status and quality of life (1). Among older adults, measurement of gait speed and time required to move from sitting to standing without using the arms, known as a chair stand, are used to evaluate physical performance, an important aspect of physical function that captures a person’s directly observed ability to conduct standardized tasks (2,3). These tests offer inexpensive, reliable measures of mobility and lower body strength and are simple to administer without specialized equipment, which make them suitable for large-scale studies and implementation in clinical practice even in resource-constrained settings (4).

In a 2018 systematic review and meta-analysis of 44 cohort studies including 101,945 individuals (mean age 72.2 years; 555 women), Veronese et. al. demonstrated that slower gait speed was a strong predictor of cardiovascular disease (CVD) and all-cause mortality (5).

However, most prior studies examining physical performance in relation to cardiovascular and all-cause mortality have not distinguished individuals with pre-existing CVD from those without it. People living with CVD typically have poorer physical function and face a higher risk of mortality than adults without a CVD history (6–8). At the same time, chronic conditions—including CVD—can contribute to functional decline, while poor physical function itself may further increase the likelihood of cardiovascular events and mortality (9). Because these pathways may differ for adults with and without established CVD, it remains uncertain whether poor physical performance continues to act as a predictor of mortality among those already affected by CVD. Clarifying potential differences in the association between physical performance and mortality between individuals with and without CVD may create opportunities to address functional limitations and potentially delay mortality.

The objective of this study was to determine if baseline history of CVD modify the association between observed physical performance, measured by gait speed and chair stand tests, and the incidence of mortality from CVD and all-causes among participants in the REasons for Geographic And Racial Difference in Stroke (REGARDS). We hypothesized that these associations would differ between participants with and without a history of CVD.

## Methods

### Study Population

The REGARDS study, a community-based prospective cohort study, enrolled 30,239 Black or White participants aged 45 years or older from the continental USA in 2003-2007. Black individuals and those living in the stroke belt or stroke buckle regions of the southeastern USA, defined by high stroke mortality rates, were over sampled (10). In 2013-2016, approximately 10 years after baseline, surviving participants were eligible for a phone interview and examination including physical performance measurement. For this analysis, we included 14,137 REGARDS participants who completed both phone and in-home interviews (**Supplementary Figure 1**).

Because of the sensitive nature of the data collected for this study, requests from qualified researchers to access the dataset may be sent to the REGARDS study at. Statistical code is available from the corresponding author.

### Physical performance measurement

Our primary exposure variables were gait speed and chair stand time. The gait speed test was conducted in participants’ homes using an 8-foot marked course; participants were instructed to walk at their usual speed while an examiner used a stopwatch to time two walks. The chair stand test required participants to complete five consecutive chair stands (standing up and sitting down without use of the arms) as quickly as possible and the time was measured in seconds. For both tests, standard procedure was followed as described in previously published studies (**11**). We excluded tests with times of less than 1 second or more than 60 seconds. Gait speed was calculated from the walk test and expressed as meter/second (m/s).

Gait speed and time to complete the chair stand test were categorized using quartiles based on the distribution in the population with and without CVD combined, with the fastest as the reference group. Participants who were unable to complete the tests were assigned to a fifth category. This is consistent with the approach used in the development of the Short Physical Performance Battery (SPPB) and allowed us to include participants with the most limited physical function who were unable to complete the tests (**2,12**).

### Outcomes

Participants were followed from the date of the in-home visit, when physical performance was assessed, through the date of mortality, loss to follow-up, or December 31, 2020. For analyses of CVD-related mortality, participants were censored if they died from other causes. Mortality was identified from multiple sources, including proxy reports during twice annual follow-up calls, the Social Security Mortality Index, and the National Death Index. Cause of mortality was adjudicated by a team of trained general internists, cardiologists or physician assistants based on interviews with the next of kin, death certificates and hospitalization records (**13**). Each death was reviewed by two independent adjudicators and if there was any disagreement, a committee decided (**13,14**).

### Covariates

History of CVD was considered present if there were any past or current instances of heart diseases, stroke, transient ischemic attack (TIA), heart failure (HF), peripheral arterial disease (PAD), deep vein thrombosis (DVT), or documented atrial fibrillation at the time physical performance was assessed based on self-reported diagnoses or adjudicated events. Socio-demographic covariates included age, sex, self-reported race, region, education level, and household income. Behavioral covariates included smoking status, alcohol consumption, and physical inactivity. In addition to history of CVD, medical characteristics included diabetes, chronic kidney disease (CKD), depressive symptoms measured using the Center for Epidemiologic Studies Depression 10-item Scale (CESD-10), perceived stress score (PSS), and systolic blood pressure (SBP). Laboratory markers included total cholesterol and high-density lipoprotein (HDL) cholesterol levels. Additionally, the use of medications such as antihypertensives, aspirin, and lipid-lowering medications were considered. Body mass index (BMI) was categorized as <25.0, ≥25.0 to <30.0, and ≥30.0 kg/m². Social support was assessed using the ENRICHD Social Support Inventory (ESSI) Score (**15**).

### Statistical Analysis

We summarized continuous variables using mean and standard deviation if normally distributed and median with interquartile range if not normally distributed. We presented categorical variables as counts and percentages by history of CVD. To address missing data and maximize the number of observations in our analysis, we employed multiple imputations by chained equations (MICE) (16). Missingness ranged from 0.02% to 28.3% across the variables; only one variable, ESSI, had >10% missing values. We conducted 20 imputations.

A priori, we conducted stratified analyses based on the history of CVD. However, we tested interaction of history of CVD with physical performance in fully-adjusted models for gait speed test and chair stand test using likelihood ratio tests. We calculated the incidence rate per 10,000 person years for CVD, and all-cause mortality by dividing the number cases by the total person-years of observation and multiplying by 10,000.

We generated Kaplan–Meier (KM) curves to visually compare survival probabilities across different levels of physical performance – gait speed and chair stand time. We examined the associations between gait speed and CVD and all-cause mortality, separately, using Cox proportional hazards regression, adjusting for potential confounding variables. Model 1 was unadjusted with only gait speed as the explanatory variable. Model 2 adjusted for sociodemographic variables, and Model 3 included all the covariates described above. The analyses were repeated for the chair stand time as the exposure variable.

Additionally, we tested possible interactions of the physical performance with sex using likelihood ratio tests. We assessed proportional hazard assumption using global Schoenfeld residual test. We also conducted sensitivity analyses constructing models with only participants with available data for all covariates (i.e., complete cases). Analyses were conducted using R (version 4.4.2; R Foundation for Statistical Computing, Vienna, Austria) and Stata (version 18; StataCorp LLC, College Station, TX). A two-sided p-value of less than 0.05 was considered statistically significant.

### Ethical approval and consent

Ethical approval for the REGARDS study was obtained from the Institutional Review Boards (IRB) at the participating institutions (UAB IRB-060217017). Written informed consent was obtained during the in-home visit.

## Results

The mean age of participants was 72.5 years, with 84% aged 65 years or older; 56.1% were women, and 36.7% were Black. At baseline, 42.9% had a history of CVD (**Table 1**). The mean age was higher among participants with a history of CVD (74.8 years) compared with those without history of CVD (70.9 years). Among participants with no history of CVD, 60.1% were women, compared with 50.7% among those with a history of CVD. A slightly higher proportion of White individuals was observed among participants with a history of CVD (64.7%) compared with those with no history of CVD (62.2%). Lower educational attainment and lower household income were more common among participants with a history of CVD. Over a median follow-up of approximately 5.9 years, there were 2,749 deaths from any cause among 14,137 participants, including 740 deaths due to CVD.

**Table 1:**
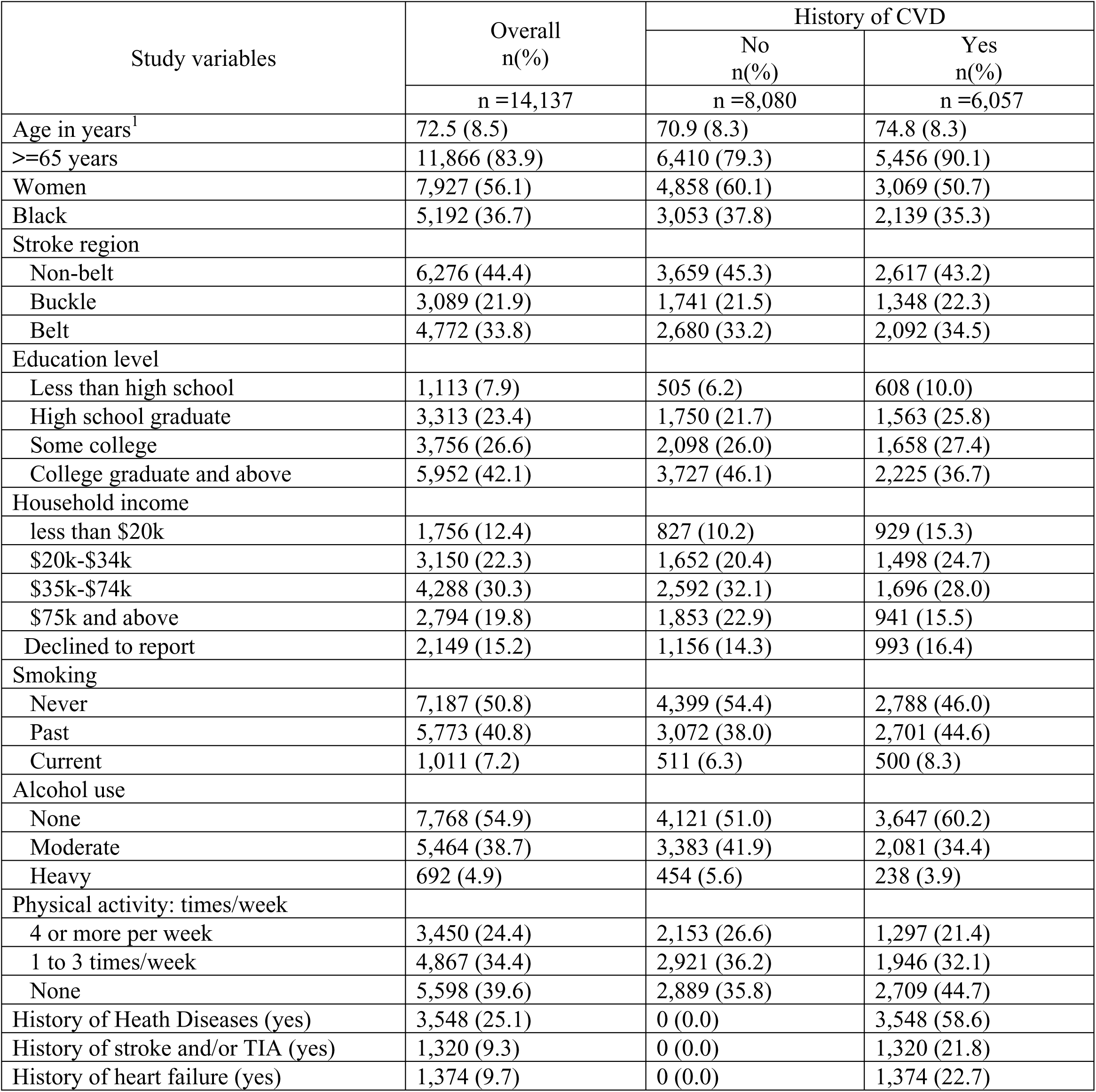
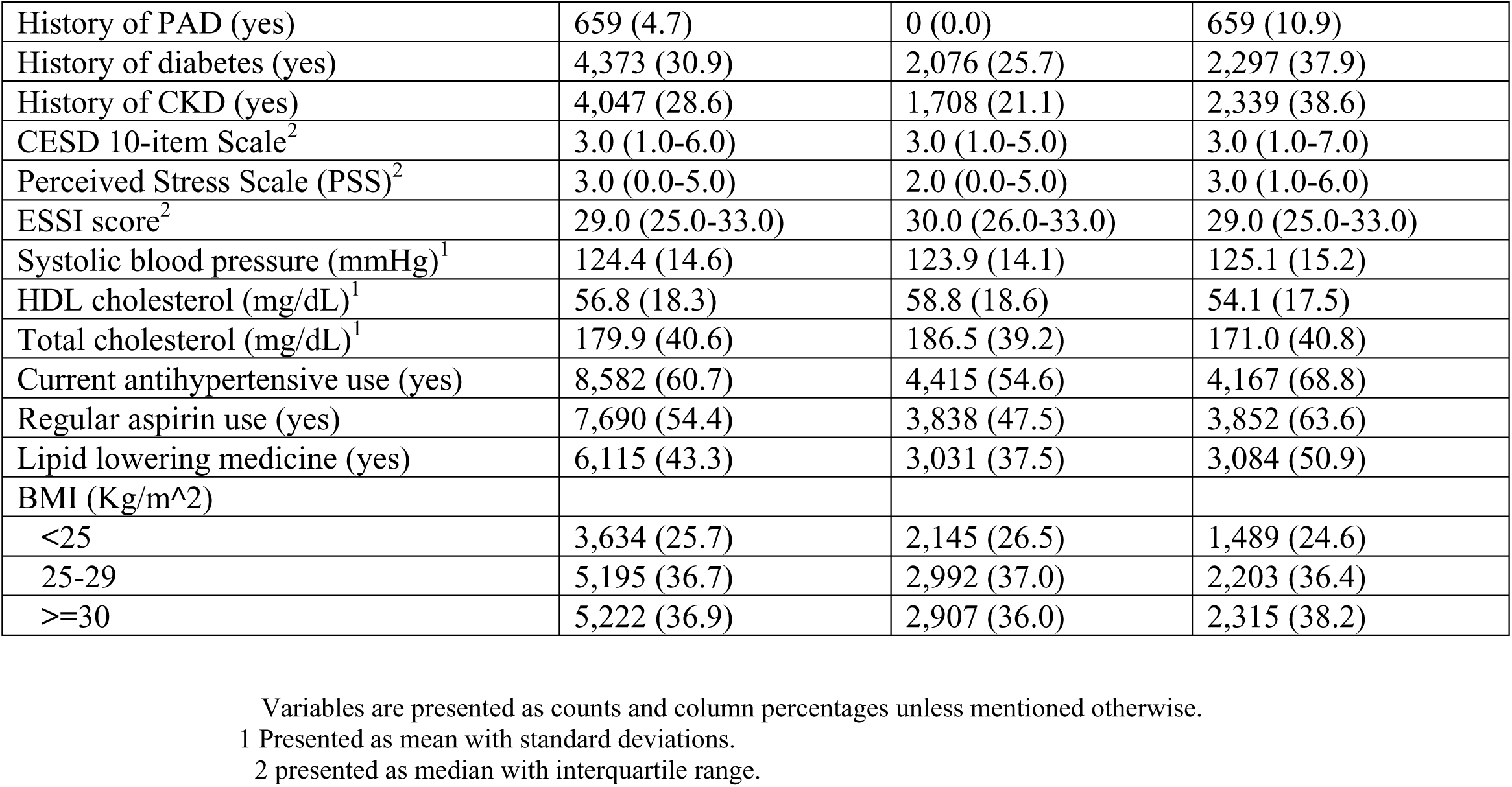
Background characteristics of the study population by history of cardiovascular diseases.

### Physical performance measurements

Among participants without history of CVD, 3.7% were unable to complete the gait speed test compared with 6.6% of those with a history of CVD; 8.9% of those without history of CVD were unable to complete the chair stand test compared with 18.0% among those with a history of CVD (**Supplementary Table 1**). Among those who completed the tests, the median gait speed was 0.61 m/s (IQR 0.37–0.86 m/s), and the median time to complete the chair stand test was 13.1 seconds (IQR 10.5–16.1).

### Association of gait speed and mortality

Among participants with and without history of CVD, survival probabilities were lower, and mortality rates were higher for both CVD and all-cause mortality among participants with slower gait speed and those who were unable to complete the timed walk tests **(Figure 1** and **Supplementary Table 1**). Among participants without history of CVD, compared with those in fastest gait speed quartile, adjusted HRs for CVD-related and all-cause mortality were 2.01 (95% CI 1.18–3.43) and 1.66 (95% CI 1.33-2.07), respectively, for participants in the lowest gait speed quartile, and 2.37 (95% CI 1.12–5.03) and 2.33 (95% CI 1.72–3.17), respectively, among those unable to complete the test (**Figure 2** and **Supplementary Table 2**). Among participants with history of CVD, compared with the fastest gait speed quartile, adjusted HRs for CVD-related and all-cause mortality were 1.28 (95% CI 0.96–1.72) and 1.72 (95% CI 1.45–2.04), respectively, for participants in the lowest gait speed quartile, and 1.87 (95% CI 1.29–2.70) and 2.74 (95% CI 2.22–3.38) among those unable to complete the test. P-values for interaction between history of CVD and gait speed were 0.028 and 0.026 for CVD-related and all-cause mortality, respectively. No significant interaction by sex was observed.

**Figure 1:**
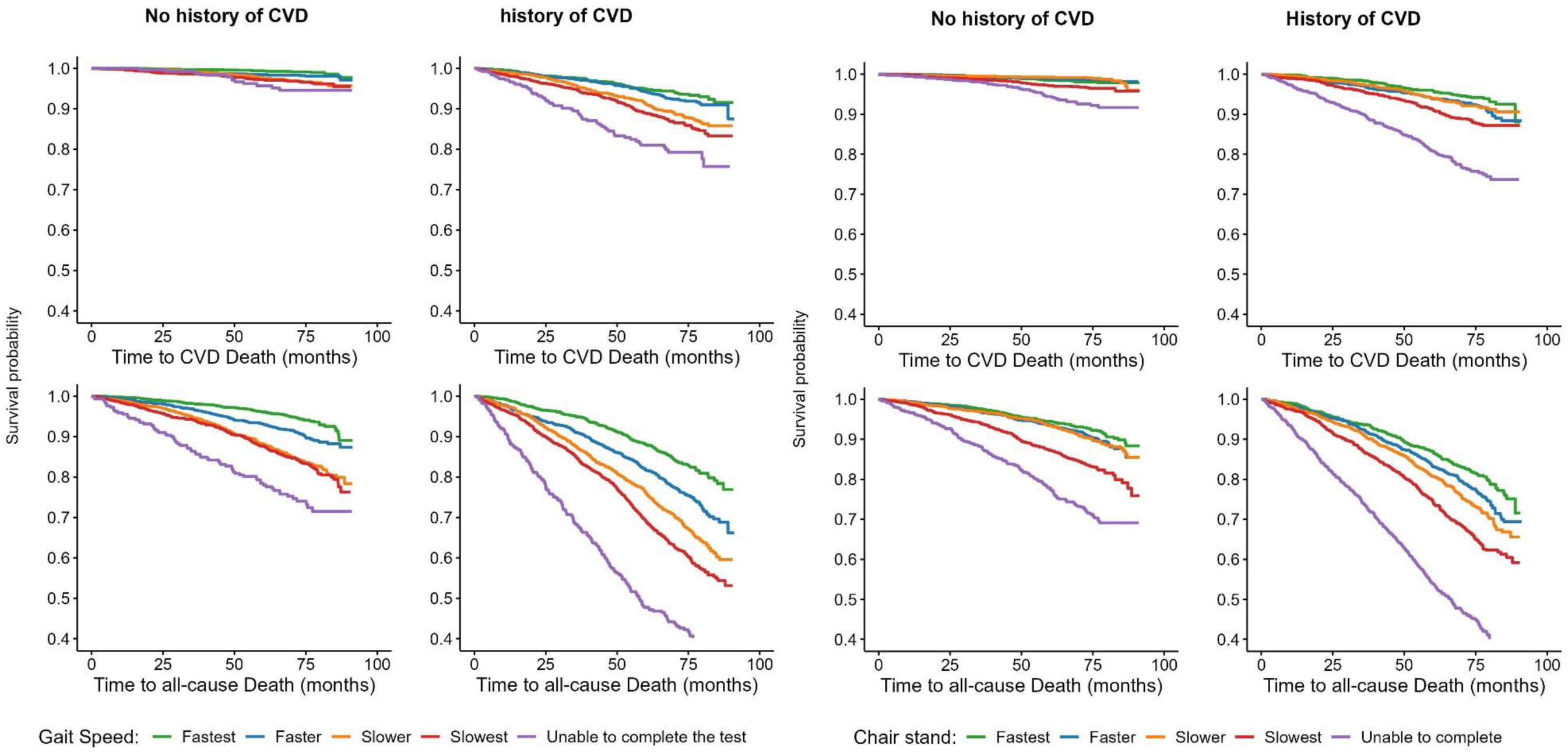
Kaplan-Meier survival curves by gait speed (Left Panel) and chair stand time (Right Panel) levels

**Figure 2:**
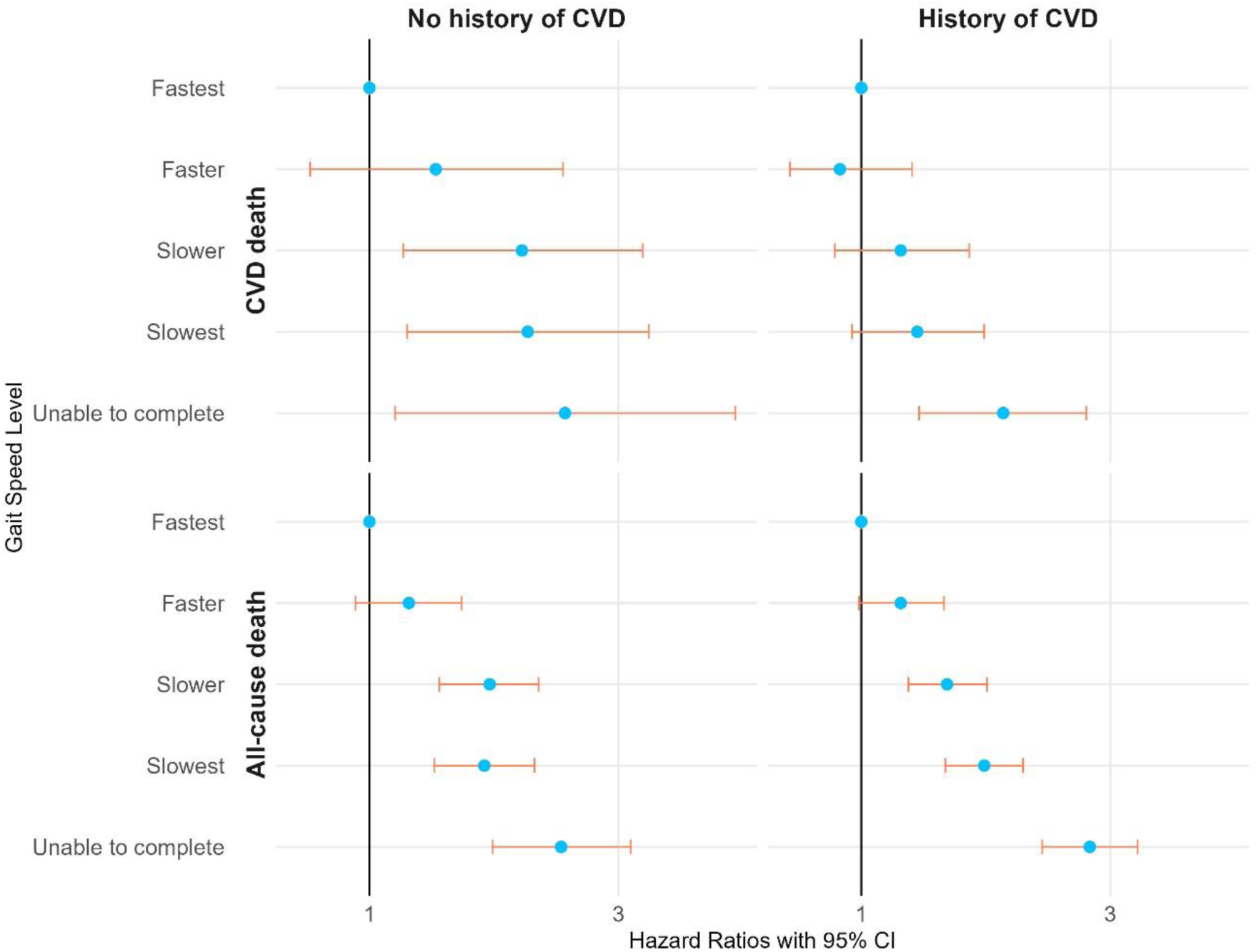
Forest plot with adjusted HRs and 95% CI for the association of CVD-related and all-cause mortality with gait speed levels [Gait Speed (m/s): Fastest >0.86; Faster >0.61–0.86; Slower >0.37–0.61; Slowest ≤0.37]. Models were adjusted for age, sex, race, region, education, income, smoking status, physical activity, depressive symptoms (CESD), systolic blood pressure, HDL cholesterol, total cholesterol, antihypertensive medication use, lipid-lowering medication use, body mass index, ESSI, and histories of heart disease, chronic kidney disease, stroke and/or transient ischemic attack, heart failure, peripheral artery disease, and diabetes.

### Association of chair stand and mortality

Among participants without history of CVD, participants in the slowest quartile of chair stand times had HRs of 0.99 (95% CI 0.62–1.56) for CVD mortality and 1.34 (95% CI 1.08–1.65) for all-cause mortality compared to the fasted quartile; those unable to complete the test had HRs of 1.68 (95% CI 1.04–2.72) and 1.82 (95% CI 1.45–2.28), respectively (**Figure 3**). Among participants with history of CVD, HRs for participants in the slowest chair stand quartile were 1.50 (95% CI 1.09–2.05) for CVD-related mortality and 1.50 (95% CI 1.26–1.78) for all-cause mortality, while those unable to complete the test had HRs of 2.29 (95% CI 1.67–3.13) and 2.23 (95% CI 1.87–2.65), respectively. P-values for interaction between history of CVD and chair stand performance were 0.084 and 0.034 for CVD-related and all-cause mortality, respectively. No significant interaction by sex was observed.

**Figure 3:**
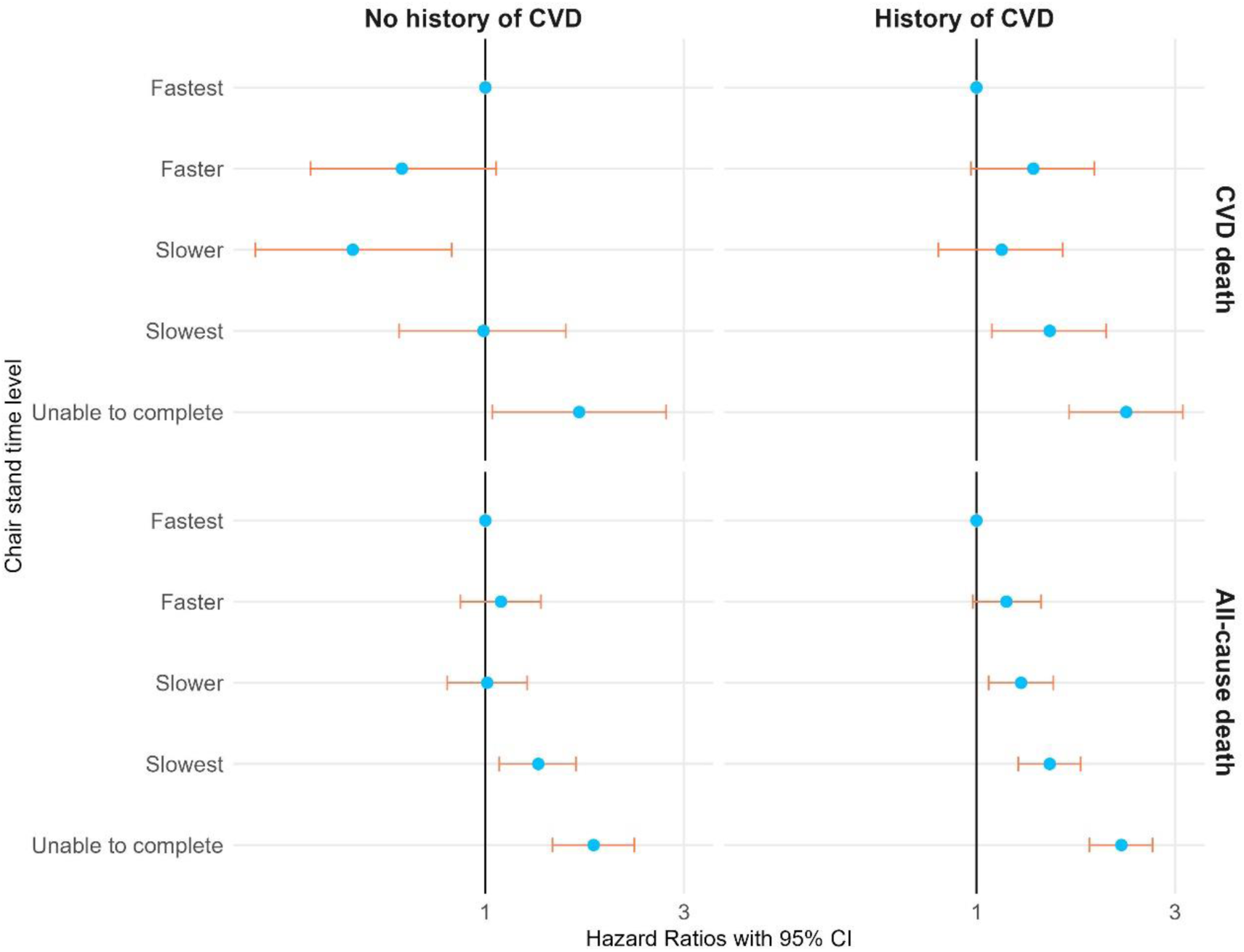
Forest plot with adjusted HRs and 95% CI for the association of CVD-related and all-cause mortality with chair stand time levels [Chair-Stand Time (seconds): Fastest ≤10.47; Faster >10.47–13.06; Slower >13.06–16.10; Slowest >16.10]. Models were adjusted for age, sex, race, region, education, income, smoking status, physical activity, depressive symptoms (CESD), systolic blood pressure, HDL cholesterol, total cholesterol, antihypertensive medication use, lipid-lowering medication use, body mass index, ESSI, and histories of heart disease, chronic kidney disease, stroke and/or transient ischemic attack, heart failure, peripheral artery disease, and diabetes.

### Sensitivity Analysis

When restricted to participants without missing data in the covariates, HRs were similar but 95% CIs were wider (**Supplementary Table 3**).

## Discussion

In this population of community-dwelling adults in the United States, slower gait speed and not being able to complete the timed walk test was associated with higher CVD-related and all-cause mortality among participants both with and without a history of CVD, though there were statistically significant differences in the magnitude of the associations by baseline history of CVD. Slower performance of the chair stands was associated with CVD and all-cause mortality among participants with history of CVD and with all-cause mortality among participants without history of CVD; being unable to complete the chair stand test was associated with CVD and all-cause mortality in both groups. These associations remained after adjustment for a wide range of sociodemographic, behavioral, and clinical variables, suggesting that physical performance captures important aspects of physiologic reserve not fully explained by traditional risk factors.

Gait speed has been broadly studied as a risk factor or predictor of CVD and all-cause mortality among older adults. Our findings align with those of Veronese et al., who conducted a meta-analysis including 44 cohort studies and more than 100,000 individuals, showing that slower gait speed is associated with higher risks of CVD and all-cause mortality (5). Similarly, Studenski et al. demonstrated that gait speed is a reliable predictor of survival among older adults, suggesting that it could be used as a clinical vital sign (17). A pooled analysis including 9 cohort studies showed that gait speed was a significant predictor of mortality and that each 0.1 m/s decrease in gait speed was associated with a 12% increase in mortality risk (17). Although less studied than gait speed, the chair stand test reflects the strength of major muscle groups in the lower body, including the quadriceps and gluteal muscles, which are essential for activities such as walking, climbing stairs, and rising from a seated position (18). Cooper et al. found that slower chair stand times were associated with higher mortality over 13 years of follow-up among British adults (19). A study by Pavasini et al. also reported that poor performance on chair stand tests was linked to increased mortality and cardiovascular events in patients with CVD (20). Muscle strength declines with age, and sarcopenia (age-related loss of muscle mass and strength) has been associated with increased morbidity and mortality (21). Our study contributes new evidence by demonstrating that poor physical performance is associated with mortality risk not only among individuals with established CVD but also among those without known CVD in a contemporary cohort of community-dwelling US adults.

The associations between gait speed and chair stand times and mortality observed in the REGARDS study and in other studies may in part be because poor physical performance is a general marker of poor health. However, physical performance is modifiable, as evidenced by studies showing that targeted interventions, such as physical therapy, physical activity, and exercise programs, can improve physical performance and reduce mortality risk (22–24).

Resistance training and exercises focusing on muscle strengthening have been shown to improve chair stand performance and overall physical function (25). Such interventions could potentially reduce mortality risks by enhancing muscle strength, improving metabolic profiles, and increasing independence in daily activities (26). The Lifestyle Interventions and Independence for Elders (LIFE) Study provides compelling evidence that physical performance is not merely a marker of health but a modifiable risk factor. This large-scale, randomized clinical trial demonstrated that a structured, moderate-intensity physical activity program significantly reduced the incidence of major mobility disability among older adults at risk for disability.

Participants in the physical activity group experienced a 30.1% incidence of major mobility disability, compared to 35.5% in the health education group over a period of 2.6 years, (HR 0.82, 95% CI 0.69-0.98; P = .03) (27). These findings underscore that interventions aimed at enhancing physical performance can lead to meaningful improvements in health outcomes, including potentially delaying mortality, thereby highlighting the importance of targeting physical performance in clinical and public health strategies.

Regular screening and monitoring of physical performance using gait speed and chair stand test may be useful to identify individuals at higher risk of adverse health outcomes and provide interventions. These simple tests can be easily implemented in various settings, including community health programs and primary care. Routine health assessments that include these measures can facilitate early detection of mobility impairments and guide timely interventions. Standardized protocols for physical performance tests can enhance consistency across studies. Evaluations of screening programs and interventions aimed at improving physical performance are needed to determine whether incorporating these measures into routine clinical and public health practice could extend life and improve function among older adults (28,29).

### Strengths and Limitations

This study benefits from several notable strengths, including the use of a large and diverse cohort from different geographic regions from the REGARDS study with detailed data collection on demographics, health behaviors, and clinical characteristics, and adjudicated cause of death. The assessment of physical performance included both gait speed and chair stand tests, which together offered assessment of mobility and strength. The study extends previous research by examining these associations among participants with and without a history of CVD. Nevertheless, there are some important limitations. The study provides only indirect evidence about whether interventions to improve physical performance would reduce mortality risk. The use of self-reported data introduces the potential for reporting bias. Despite adjusting for numerous covariates, residual confounding from unmeasured or incompletely measured characteristics such as nutritional status, unrecognized cognitive impairment, subclinical disease (e.g., early heart failure or neurodegenerative conditions), chronic inflammation, and psychosocial factors such as depression or social isolation cannot be ruled out. Physical performance was measured at a single time point; we were not able to determine whether physical activity and comorbidities were causes of physical performance, and therefore confounders, or results of physical performance, and therefore on the causal pathway between physical performance and mortality. Gait speed was measured using an 8-foot timed walk because this distance was considered to be practical to implement in participants’ homes.

However, gait speed is systematically lower when measured using short walk courses, compared to longer distances (30). Furthermore, the study’s generalizability is limited, as it included community-dwelling older adults, excluding individuals living in nursing homes and those with severe disabilities. Also, significant population groups in the US including Asian Americans and Hispanic Americans—were not included, which may limit the applicability of the results to these populations.

### Conclusion

Poor physical performance was associated with higher CVD-related and all-cause mortality among individuals both with and without a history of CVD. Although mortality rates were higher among those with history of CVD, the relative associations between physical performance and mortality were substantial in the group without a history of CVD, indicating that performance-based assessments may identify adults at elevated risk even in the absence of CVD.

## Acknowledgement

The REGARDS study is supported by cooperative agreement U01 NS041588 co-funded by the National Institute of Neurological Disorders and Stroke (NINDS) and the National Institute on Aging (NIA), National Institutes of Health, Department of Health and Human Service. The content is solely the responsibility of the authors and does not necessarily represent the official views of the NINDS or the NIA. Representatives of the NINDS were involved in the review of the manuscript but were not directly involved in the collection, management, analysis or interpretation of the data. Additional funding for this work was provided by R01 HL80477 and R01 HL165452 from the National Heart Lung and Blood Institute (NHLBI). The authors thank the other investigators, the staff, and the participants of the REGARDS study for their valuable contributions. A full list of participating REGARDS investigators and institutions can be found at: https://www.uab.edu/soph/regardsstudy/

## Supplementary tables and figures

**Supplementary Figure 1:**
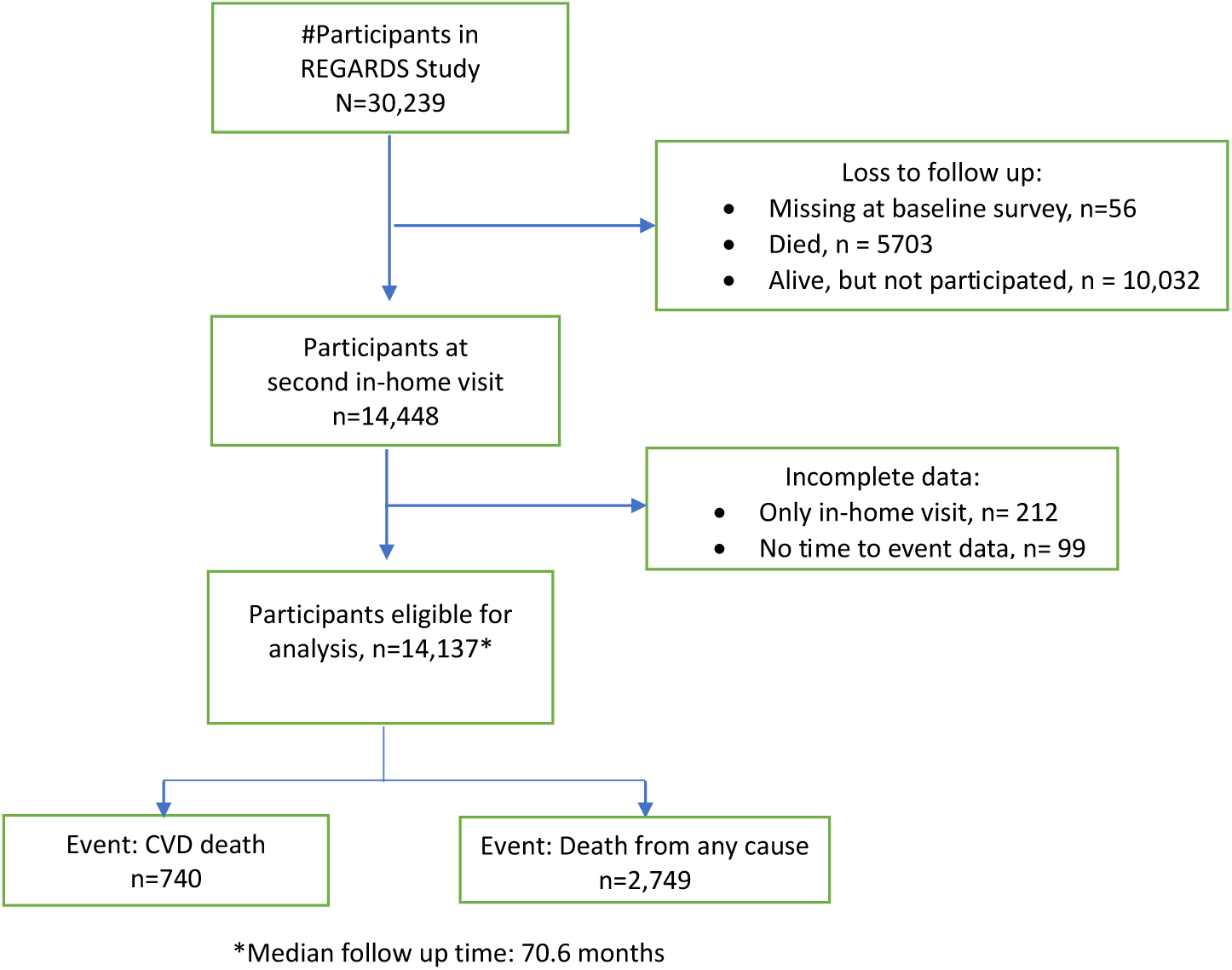
Participants selection flowchart.

**Supplementary table 1:**
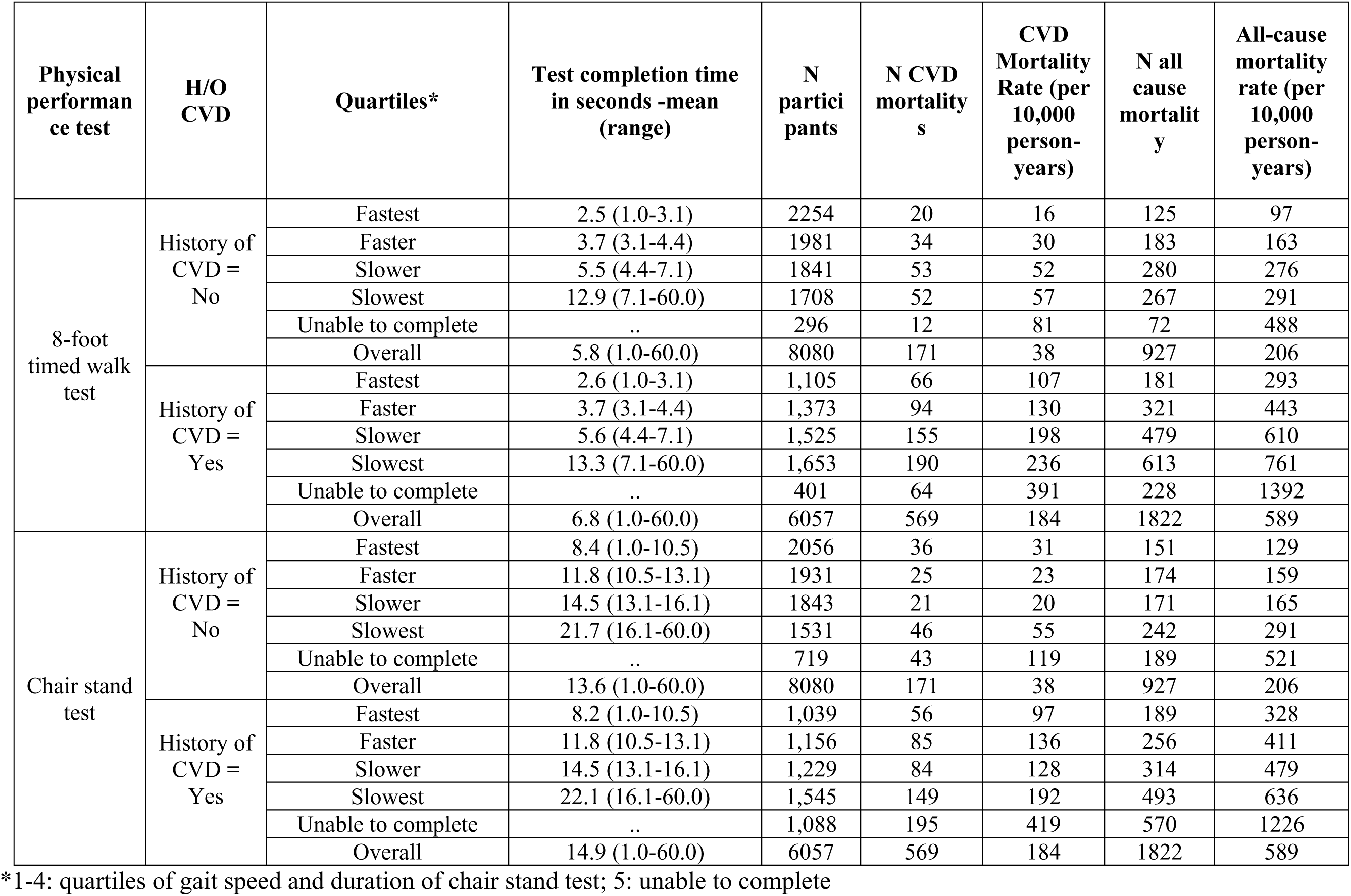
Summary of Physical Performance Measures and CVD and all-cause mortality by history of CVD.

**Supplementary Table 2:**
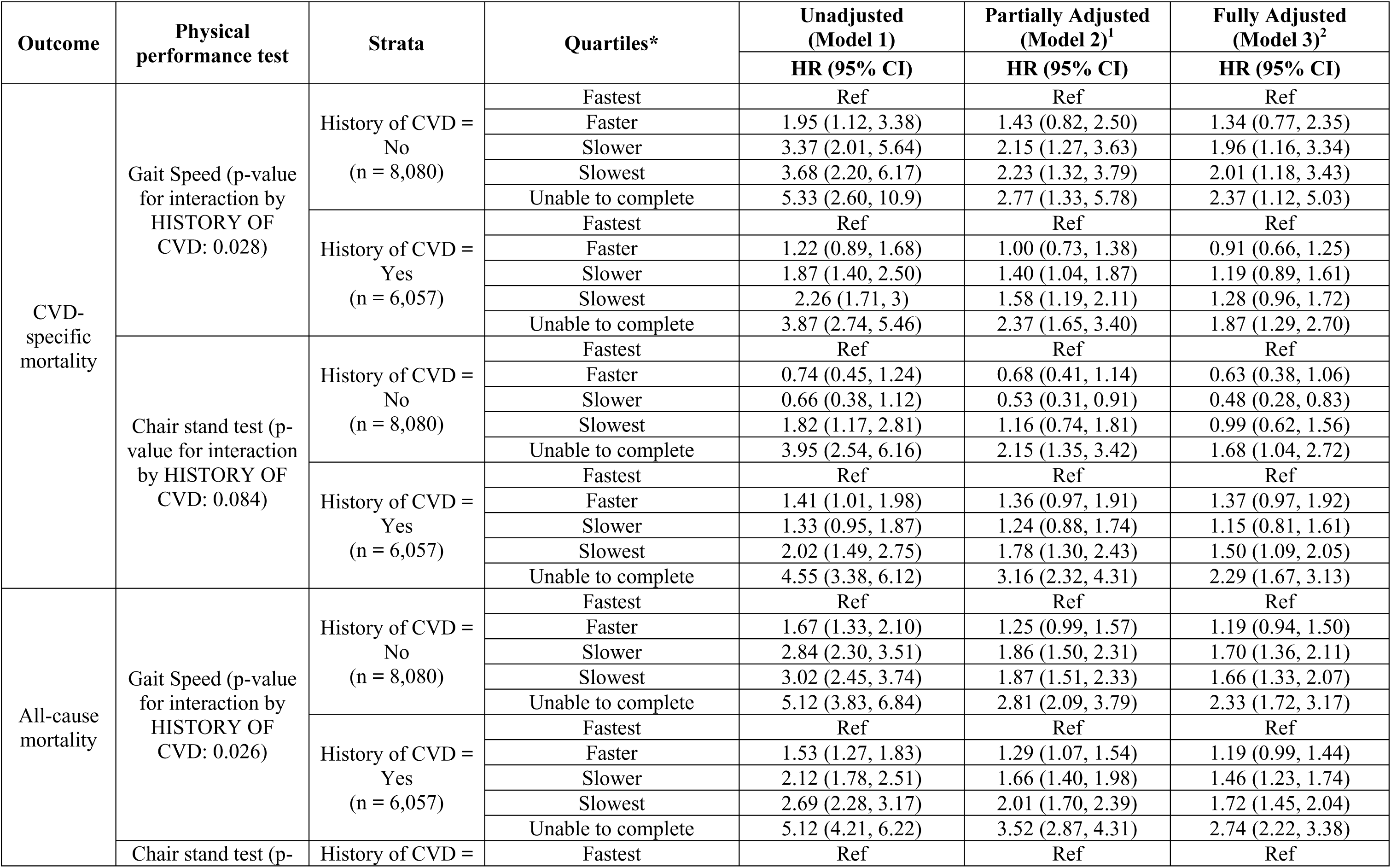
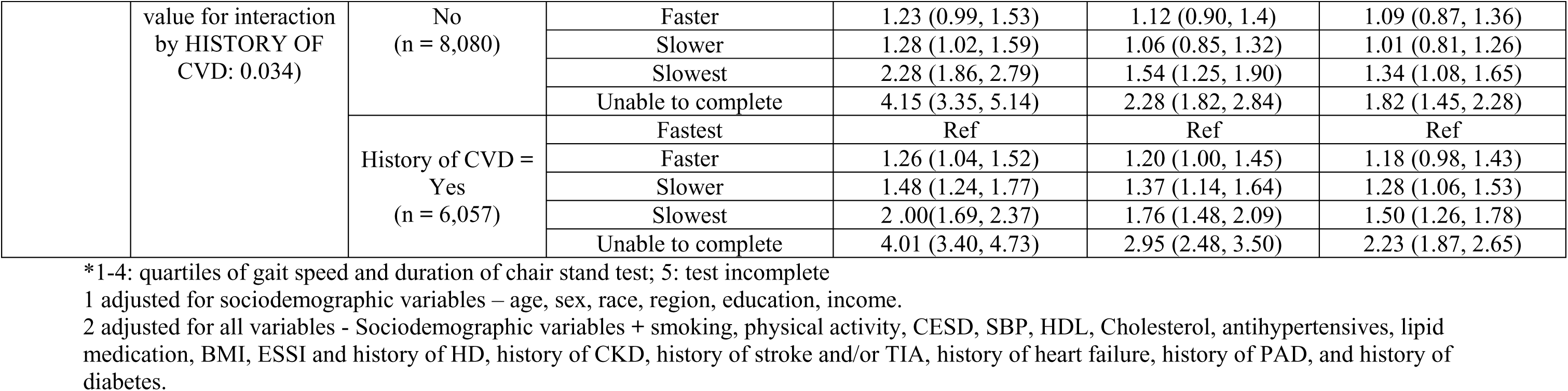
Association Between Physical Performance and CVD-specific and all-cause mortality*.

**Supplementary table 3:** Association Between Physical Performance and CVD-specific and all-cause mortality: Complete Cases vs. Imputed Data.

| Outcome | Physical performance test | Strata | Quartiles | Full models with complete cases | Full models with imputed data |
| --- | --- | --- | --- | --- | --- |
|  |  |  |  | HR (95% CI) | HR (95% CI) |
| CVD | Gait Speed | History of CVD = No | Fastest | Ref | Ref |
|  |  |  | Faster | 0.98 (0.54, 1.78) | 1.34 (0.77, 2.35) |
|  |  |  | Slower | 1.08 (0.60, 1.93) | 1.96 (1.16, 3.34) |
|  |  |  | Slowest | 1.41 (0.81, 2.46) | 2.01 (1.18, 3.43) |
|  |  |  | Unable to complete | 3.49 (1.85, 6.59) | 2.37 (1.12, 5.03) |
|  |  | History of CVD = Yes | Fastest | Ref | Ref |
|  |  |  | Faster | 1.00 (0.73, 1.38) | 0.91 (0.66, 1.25) |
|  |  |  | Slower | 1.40 (1.04, 1.87) | 1.19 (0.89, 1.61) |
|  |  |  | Slowest | 1.58 (1.19, 2.11) | 1.28 (0.96, 1.72) |
|  |  |  | Unable to complete | 2.37 (1.65, 3.40) | 1.87 (1.29, 2.70) |
|  | Chair stand test | History of CVD = No | Fastest | Ref | Ref |
|  |  |  | Faster | 0.99 (0.42, 2.32) | 0.63 (0.38, 1.06) |
|  |  |  | Slower | 0.50 (0.18, 1.40) | 0.48 (0.28, 0.83) |
|  |  |  | Slowest | 0.91 (0.35, 2.35) | 0.99 (0.62, 1.56) |
|  |  |  | Unable to complete | 2.39 (0.88, 6.49) | 1.68 (1.04, 2.72) |
|  |  | History of CVD = Yes | Fastest | Ref | Ref |
|  |  |  | Faster | 1.20 (0.77, 1.87) | 1.37 (0.97, 1.92) |
|  |  |  | Slower | 0.99 (0.63, 1.55) | 1.15 (0.81, 1.61) |
|  |  |  | Slowest | 1.28 (0.84, 1.94) | 1.50 (1.09, 2.05) |
|  |  |  | Unable to complete | 2.11 (1.39, 3.20) | 2.29 (1.67, 3.13) |
| All cause | Gait Speed | History of CVD = No | Fastest | Ref | Ref |
|  |  |  | Faster | 1.10 (0.82, 1.48) | 1.19 (0.94, 1.50) |
|  |  |  | Slower | 1.66 (1.26, 2.20) | 1.70 (1.36, 2.11) |
|  |  |  | Slowest | 1.62 (1.22, 2.16) | 1.66 (1.33, 2.07) |
|  |  |  | Unable to complete | 1.30 (0.75, 2.26) | 2.33 (1.72, 3.17) |
|  |  | History of CVD = Yes | Fastest | Ref | Ref |
|  |  |  | Faster | 1.44 (1.08, 1.92) | 1.19 (0.99, 1.44) |
|  |  |  | Slower | 1.59 (1.19, 2.12) | 1.46 (1.23, 1.74) |
|  |  |  | Slowest | 1.92 (1.45, 2.55) | 1.72 (1.45, 2.04) |
|  |  |  | Unable to complete | 3.09 (2.12, 4.50) | 2.74 (2.22, 3.38) |
|  | Chair stand test | History of CVD =<br>No | Fastest | Ref | Ref |
|  |  |  | Faster | 1.25 (0.94, 1.66) | 1.09 (0.87, 1.36) |
|  |  |  | Slower | 0.93 (0.69, 1.26) | 1.01 (0.81, 1.26) |
|  |  |  | Slowest | 1.45 (1.09, 1.92) | 1.34 (1.08, 1.65) |
|  |  |  | Unable to complete | 1.56 (1.11, 2.19) | 1.82 (1.45, 2.28) |
|  |  | History of CVD =<br>Yes | Fastest | Ref | Ref |
|  |  |  | Faster | 1.23 (0.95, 1.58) | 1.18 (0.98, 1.43) |
|  |  |  | Slower | 1.33 (1.04, 1.70) | 1.28 (1.06, 1.53) |
|  |  |  | Slowest | 1.45 (1.15, 1.84) | 1.50 (1.26, 1.78) |
|  |  |  | Unable to complete | 2.14 (1.67, 2.75) | 2.23 (1.87, 2.65) |

